# Effect of gastroesophageal reflux disease on sleep disorders: a Mendelian randomization study

**DOI:** 10.1101/2023.02.11.23285798

**Authors:** Zijie Li, Weitao Zhuang, Junhan Wu, Haijie Xu, Yong Tang, Guibin Qiao

**Affiliations:** Department of Thoracic Surgery, Guangdong Provincial People’s Hospital (Guangdong Academy of Medical Sciences), Southern Medical University, Guangzhou 510080, China; Shantou University Medical College, Shantou 515041, China; Department of Medical Oncology, State Key Laboratory of Oncology in South China, Collaborative Innovation Center for Cancer Medicine, Sun Yat-sen University Cancer Center, Guangzhou 510060, China; Department of Thoracic Surgery, The First Affiliated Hospital of Shantou University Medical College, Shantou 515041, China

**Keywords:** gastroesophageal reflux disease, sleep disorders, sleep apnoea, Mendelian randomization

## Abstract

**Background:** Recently observational studies have consistently shown an association between gastroesophageal reflux disease (GERD) and sleep disorders. In this study, Mendelian randomization (MR) analysis was performed to determine the genetic causal relationship between GERD and the risk of sleep disorders.

**Methods:** The summary statistics of GERD and sleep disorders were obtained through large-scale genome-wide association studies (GWAS). In addition to exploring sleep disorders, a deeper analysis was conducted on some major categories of sleep disorders such as sleep apnoea and insomnia. Various MR analysis methods including inverse-variance weighted (IVW), MR Egger, weighted median, simple mode, and weighted mode were performed, and the results of IVW were taken as the primary results. In addition, sensitivity analyses including heterogeneity test and pleiotropy test were also performed to test the robustness of the MR results.

**Results:** After removing the ineligible SNPs and the palindromic SNPs, IVW detected a significant effect of GERD on sleep disorders (OR = 1.436, 95% CI: 1.309-1.576, p = 2.099E-14) and sleep apnoea (OR = 1.486, 95% CI: 1.341-1.647, p = 4.409E-14). However, there was no genetic causality in the effect of GERD on insomnia (IVW OR = 1.146, 95% CI: 0.877-1.498, p = 0.319). Furthermore, the heterogeneity test and pleiotropy test found no evidence of bias, which indicated the results were robust.

**Conclusions:** Our study found that the presence of GERD increased the risk of sleep disorders and sleep apnoea, but not the risk of insomnia. Further research is needed to identify the specific mechanisms of causal relationships.

## Background

Gastroesophageal reflux disease (GERD) is an illness describing the reflux of gastric contents into the esophagus. Acid regurgitation and heartburn are common uncomfortable symptoms of GERD [1]. Long-term GERD can lead to esophagitis and also increase the risk of esophageal cancer [1]. Approximately 13% of the general population worldwide suffer from GERD at least once a week [2].

Besides the esophageal symptoms, GERD has numerous extra-esophageal symptoms, among which chronic cough and laryngitis are two well-known clinical manifestations [3]. Remarkably, it has been indicated that there might be a strong correlation between GERD and sleep disorders in many previous studies [4, 5], and the relationship might be bidirectional [6-9]. GERD not only affects the objective sleep quality, but also may lead to the development of obstructive sleep apnoea (OAS) and insomnia [4, 10]. On the one hand, previous studies have shown that therapy for GERD could impair subjective sleep parameters and improve sleep quality in patients with sleep disorders [4, 6, 11]. What is more, continuous positive airway pressure treatment in patients with OSA has been shown to simultaneously improve sleep quality and mitigate acid regurgitation [12].

There has been controversy regarding the bidirectional relationship between GERD and sleep disorders which needs to be urgently resolved. Recently, a study has genetically confirmed that sleep disorders such as insomnia and short sleep are risk factors for GERD [13]. However, the essential relationship between GERD and the risk of developing sleep disorders has not been robustly demonstrated. Moreover, existing observational epidemiological findings may be influenced by confounding factors and reverse causality, making causal inference difficult [14]. It is of great significance to demonstrate the causal relationship between GERD and sleep disorders, which is particularly relevant for the adjustment of clinical interventions.

It is impractical and unethical to evaluate the effect of GERD on sleep disorders by randomized controlled trials (RCTs), even though they are the best causal inference method in etiology. Mendelian randomization (MR) analysis uses single nucleotide polymorphisms (SNPs) as proxies of exposure to assess the effect on outcome [15, 16]. Genetic instrumental variables (IVs) are assumed to affect outcome only through exposure, without confounding factors. Therefore, MR analysis is less influenced by reverse causality and confounders [17]. This study aims to explore the potential causal association between genetic liability for GERD and the risk of sleep disorders through MR analysis.

## Methods

### Study design

Our study followed the newly generated STROBE-MR statement for reporting MR research [18]. The basic process of MR analysis was shown in Figure 1. In this study, we set GERD and sleep disorders as exposure and outcome, respectively. The genetic information of GERD derived from a genome-wide association study (GWAS) was set as IVs.

**Figure 1.**
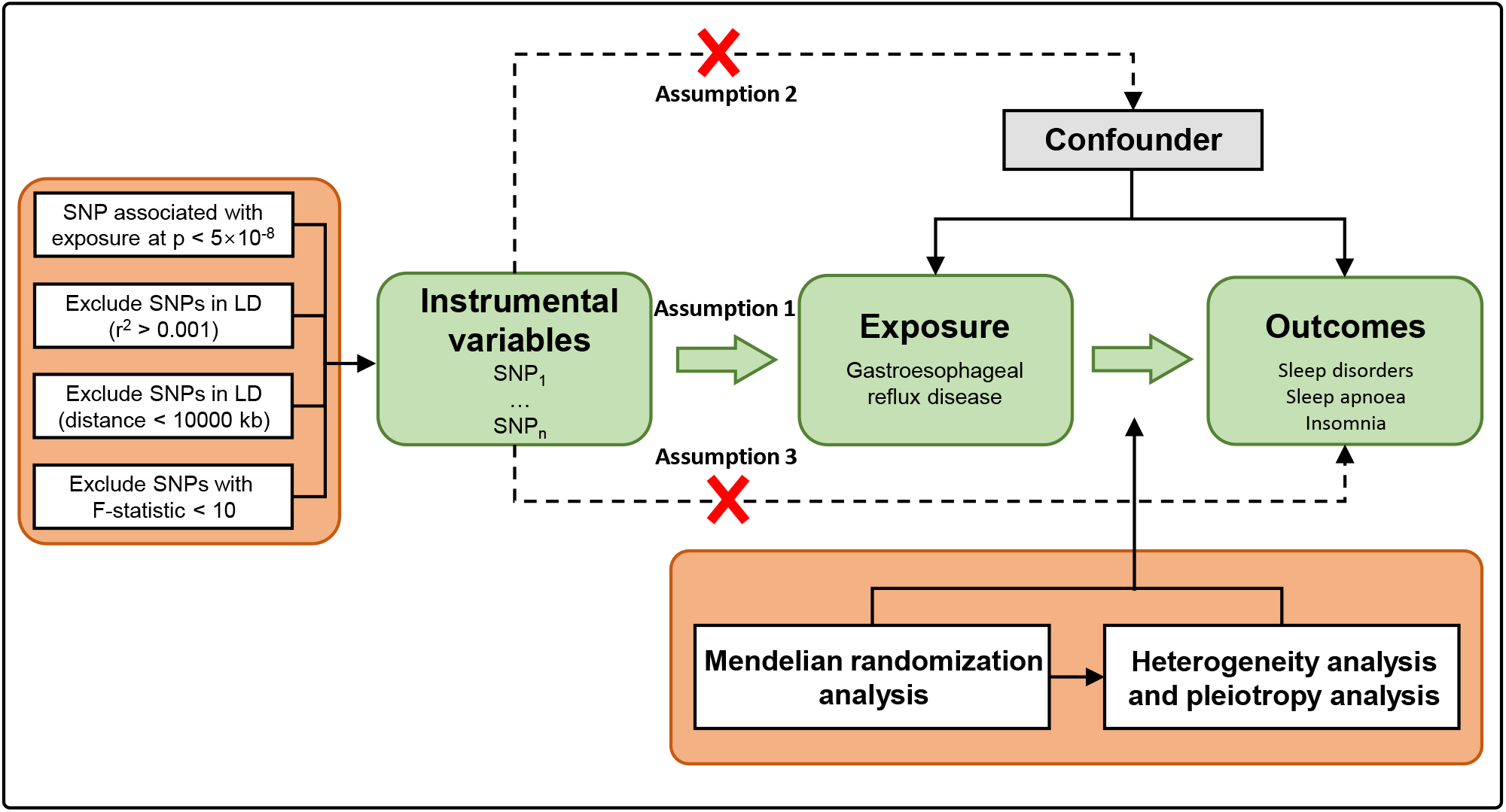
Flow chart of this study. SNPs, single-nucleotide polymorphisms; LD, linkage disequilibrium.

### Data sources

For GERD, we retrieved the GWAS summary data from the largest published GWAS study of GERD in European populations, which included 129,080 cases and 473,524 controls [19]. For sleep disorders, there are seven main categories of sleep disorders [20]. Previous observational studies have shown that GERD has an impact on sleep disorders, particularly on sleep apnoea and insomnia [4, 10]. Therefore, in addition to exploring sleep disorders, we have also analyzed several major categories of sleep disorders such as sleep apnoea and insomnia for a deeper analysis. GWAS summary statistics for several aforementioned phenotypes were obtained from the FinnGen Consortium (Table 1). All the GWAS summary data applied in this study can be obtained from the IEU open GWAS project (https://gwas.mrcieu.ac.uk/).

**Table 1.** Basic information on the GWAS applied in this study.

| Year | Trait | Consortium | Population | Sample size | N-case | N-control | Number of SNPs |
| --- | --- | --- | --- | --- | --- | --- | --- |
| 2021 | Gastroesophageal reflux<br>disease | PMID: 34187846 | European | 602,604 | 129,080 | 473,524 | 2,320,781 |
| 2021 | sleep disorders | FinnGen | European | 216,700 | 19,155 | 197,545 | 16,380,458 |
| 2021 | sleep apnoea | FinnGen | European | 217,955 | 16,761 | 201,194 | 16,380,465 |
| 2021 | insomnia | FinnGen | European | 217,855 | 1,691 | 216,164 | 9,851,867 |

SNPs, Single nucleotide polymorphisms.

### Selection of IVs

MR is based on three critical assumptions: (1) IVs are closely related to GERD; (2) IVs should not be affected by known or unknown confounding factors; and (3) IVs only affect sleep disorders through GERD [21]. The following standard procedures were used to select the best IVs, which ensured the integrity and accuracy of the results. First, SNPs that reached the genome-wide significant threshold (p < 5 × 10^−8^) were extracted. To ensure the independence of IVs, a clumping process (r^2^ < 0.001 and distance > 10,000 kb) was performed, which used linkage disequilibrium (LD) estimates calculated from Europeans in the 1000 Genomes project [22]. Second, ensuring that there is no association between the instrumental SNPs and confounding factors, which was identified through the PhenoScanner database [23]. Third, the GERD SNPs that were closely associated (p < 5 × 10^−5^) with sleep disorders were considered ineligible SNPs and were further excluded. Furthermore, F-statistics is the measurement of the strength of IVs, which equals to R^2^ × (N − 2) ÷ (1 − R^2^), where R^2^ refers to the proportion of exposure variability explained by each IV and N refers to the sample size of the GWAS for exposure [24]. R^2^ is calculated by the equation: 2 × EAF × (1 − EAF) × Beta^2^, which used to represent the proportion of variance in an exposure factor explained by the IVs. EAF is the effect allele frequency and Beta represents the estimated genetic effect on the risk of GERD [25]. F-statistics > 10 are typically used as the cutoff for powerful IVs, and F-statistics less than 10 usually indicate the weak instrument bias [26].

### MR analysis methods

In this study, the inverse variance weighted (IVW) method was conducted as the primary analysis method to identify significant causal associations between genetic liability to GERD and the risk of sleep disorders with p <□0.05. In addition, we performed the following additional methods, including MR Egger, weighted median, simple mode, and weighted mode, which enhanced the reliability of causal results [27]. If the estimates direction of the above MR methods were significantly similar, it indicated that the causal effect of GERD on sleep disorders was stable and reliable.

### Sensitivity analysis

The Cochrane’s Q test was used to assess heterogeneity between SNPs in the IVW method. It was considered that there was no heterogeneity in the causal analysis when the p-value□≥□0.05 [28]. Meanwhile, the random-effects IVW test was utilized to provide a more conservative yet robust estimate when heterogeneity exists (p < 0.05). The MR Pleiotropy RESidual Sum and Outlier (MR-PRESSO) test was applied to detect potential horizontal pleiotropy and correct it by removing outlier SNPs [29]. In addition, the leave-one-out analysis was performed to evaluate the reliability of the affiliation between the SNPs and GERD, evaluating whether any SNP was responsible for the significant results. The funnel plot was also used to detect the heterogeneity, and a symmetry plot indicated the absence of heterogeneity. The results are assumed to be robust when the heterogeneity and pleiotropy of the results do not exist. All statistical analyses were performed in R software v4.0.0 (R Core Team 2020) with the “TwoSampleMR” v0.6.0 package, and the “MRPRESSO” v1.0 package. The forest plot figure was generated in the “forestplot” v3.11 package.

## Results

### MR analysis

After removing the ineligible SNPs and the palindromic SNPs, the remaining SNPs were used for the subsequent MR analysis (Supplemental Table 1-3). All the F-statistics were greater than 10 (ranging from 208 to 669), indicating no evidence of weak instrument bias. As shown in figure 2, the IVW analysis exhibited an increased risk of sleep disorders in patients with GERD (p = 2.099E-14). Meanwhile, the estimates from MR Egger (p = 0.237), weighted median (p = 3.629E-09), simple mode (p = 8.889E-03), and weighted mode (p = 5.209E-03) showed a consistent direction of the IVW estimate. In addition, IVW analysis also observed a causal relationship between GERD and sleep apnoea (p = 4.409E-14), and other MR analysis methods were also statistically consistent. However, there is no causal relationship between GERD and insomnia (IVW OR = 1.146, 95% CI: 0.877-1.498, p = 0.319).

**Figure 2.**
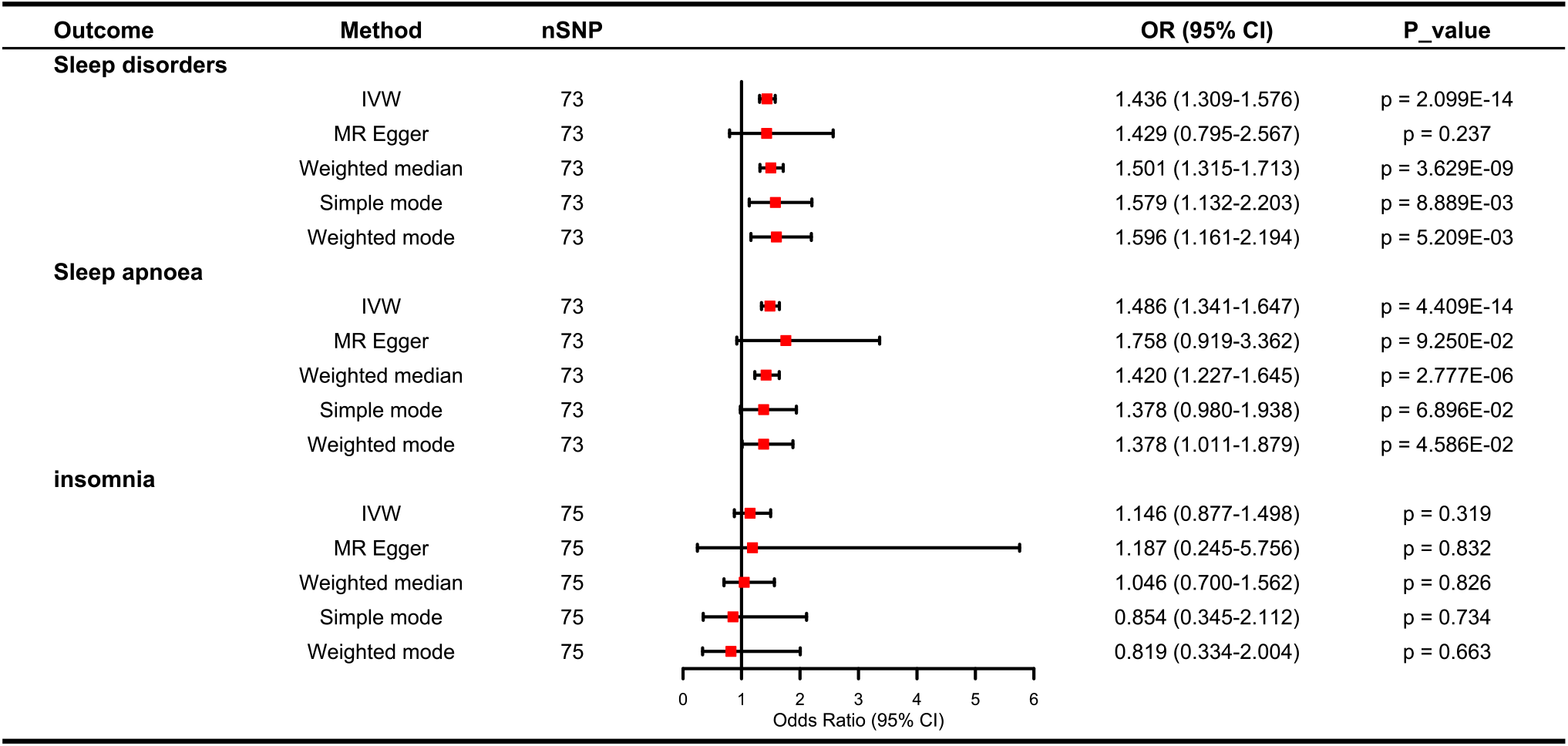
Mendelian randomization estimates of the associations between GERD and the risk of sleep disorders. IVW, Inverse variance weighted。

### Sensitivity analyses

According to the result of Cochran’s Q test, there was no heterogeneity observed in MR Egger and IVW (Table 2). The Egger-intercepts of MR-Egger analyses were not statistically significantly different from zero (p = 0. 986, p = 0. 609, and p = 0. 965), and directional horizontal pleiotropy was not detected, which meant the SNPs of GERD do not affect the incidence of sleep disorders, sleep apnoea, and insomnia through traits other than GERD. The MR-PRESSO test did not detect any horizontal pleiotropy among the instrumental SNPs either (Table 2).

**Table 2.** Heterogeneity pleiotropy test of GERD genetic IVs for sleep disorders.

| Heterogeneity test | Pleiotropy test |
| --- | --- |

| Outcomes | MR Egger |  |  | Inverse variance weighted |  |  | MR Egger |  |  | PRESSO |
| --- | --- | --- | --- | --- | --- | --- | --- | --- | --- | --- |
|  | Q | Q_df | P value* | Q | Q_df | P value* | intercept | SE | P value* | P value* |
| sleep disorders | 74.49 | 71 | 0.366 | 74.49 | 72 | 0.397 | 0.0001 | 0.009 | 0.986 | 0.456 |
| sleep apnoea | 79.27 | 71 | 0.234 | 79.57 | 72 | 0.253 | -0.0055 | 0.011 | 0.609 | 0.310 |
| insomnia | 60.62 | 73 | 0.849 | 60.63 | 74 | 0.868 | -0.0012 | 0.026 | 0.965 | 0.867 |

\*P value $\geq 0.05$ represents no significant heterogeneity or pleiotropy.

MR regression slopes showed positive correlations between the effect of SNP on GERD and sleep disorders as well as sleep apnoea (Figure 3). The results of the leave-one-out analysis demonstrated that the risk estimates of genetically predicted GERD on sleep disorders, sleep apnoea, and insomnia were remarkably stable after leaving out one SNP at a time (Supplemental Figure 1). Moreover, the forest plots and funnel plots also showed that there was no significant heterogeneity of the instrumental variable SNPs (Supplemental Figure 2,3)

**Figure 3.**
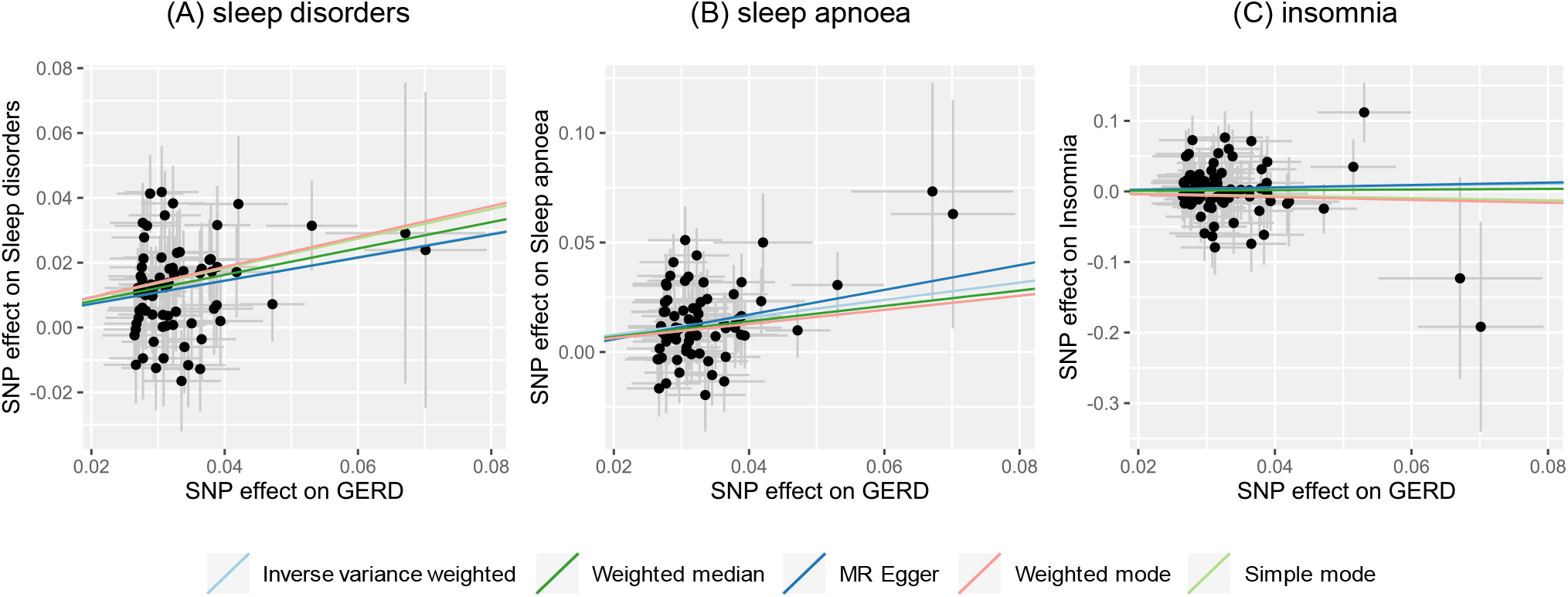
Scatter plots of the effect size of each SNP on GERD and sleep disorders. (A) sleep disorders, (B) sleep apnoea, (C) insomnia. GERD, Gastroesophageal reflux disease.

## Discussion

It is challenging to confirm the causal relationship between GERD and the risk of sleep disorders without long-term prospective studies and RCTs. Although previous observational studies have found a high prevalence of sleep disorders in patients with GERD, the results of these studies are susceptible to confounders as well as reverse causation, which may affect the robustness of the inferred conclusions. This study provided a genetically potential causal proof that patients with GERD have a greater risk of sleep disorders and sleep apnoea, which strongly confirmed the results of previous observational studies and contributed to clinical strategy optimization.

Sleep disorders as well as sleep apnoea may be primarily associated with nocturnal GERD rather than daytime GERD. A previous large-scale observational study has shown that nocturnal gastroesophageal reflux symptom and sleep disorders were both highly prevalent among patients with GERD (88.9% and 68.3%, respectively). Patients with nocturnal GERD symptoms were 50% more likely to suffer from sleep disorders than those with GERD symptoms only during the day [5]. Similarly, Ossur Ingi Emilsson et al. also found that the patients with nocturnal GERD had more OSA symptoms than the patients without nocturnal GERD [30]. Furthermore, the treatment of nocturnal GERD including proton-pump inhibitors has been shown to improve both subjective and objective sleep measurements [8, 11]. The above evidence revealed that nocturnal gastroesophageal reflux symptoms may be a major risk factor for sleep disorders in patients with GERD.

GERD symptoms negatively affect the quality of sleep and cause sleep disorders. Kindt S et al. found that 84% of reported sleep disorders were attributed to nocturnal GERD, and the main causes of sleep disorders due to GERD included reflux in the supine position (72%), typical reflux disturbing sleep (39%), and waking because of reflux (45%), among 9322 patients with heartburn or regurgitation [31]. In addition, Jui-Sheng Hung et al. indicated that increased nocturnal acid reflux may play a role in inducing sleep disorders in patients with GERD [32]. Normally, gravity and peristaltic clearance of esophageal reflux can limit the contact time between esophageal acid and mucosa. In contrast, during recumbent sleep, the beneficial effect of gravity is lost. Meanwhile, salivation and swallowing are dramatically decreased and peristalsis of esophageal wall is repressed, especially during deep sleep, which can lead to prolonged acid contact in the esophagus and favor upward migration of refluxate into the proximal esophagus during sleep [33]. When gastroesophageal reflux occurred during sleep, it might cause heartburn sensations, which may interfere with sleep and lead to sleep disorders. Moreover, the severity of sleep disorders is closely related to the severity of nocturnal acid reflux and esophageal pH changes [34]. Furthermore, a significant proportion of people with GERD are more likely to have accompanying conditions such as functional dyspepsia and irritable bowel syndrome, which affected the quality of life and might have an impact on sleep disorders [35].

Notably, multiple mechanisms may exist between GERD and the high risk of sleep apnoea, mainly including the vagal reflex, and the repetitive microaspiration of refluxed gastric acid by the respiratory tract. Most patients with GERD have nocturnal acid reflux symptoms because the lying position during sleep can lead to the migration of gastric acid and aspiration into the respiratory tract [36]. During the sleep period, the refluxed gastric acid in the distal esophagus might trigger the vagal reflex, which facilitates bronchospasm, vasodilatation and mucus secretion, resulting in airway stenosis and obstruction [37]. The migration of refluxed gastric contents and microaspiration of acid also could cause inflammation and edema of the upper airway, as well as bronchoconstriction, thereby predisposing to OSA.

This was the first MR study to illustrate the causal relationship between GERD and the high risk of sleep disorders. The current study had several strengths. First, the MR study is different from the observational study in that genetic associations can be obtained from large-scale GWAS, which can significantly improve the statistical efficacy of causal relationship. Second, the MR study is not susceptible to confounding factors and reverse causality that are commonly seen in observational studies. Third, the sensitivity analysis including the heterogeneity test and pleiotropy test can verify the robustness of the results, thereby strengthening the evidence for our findings.

Despite the validity and robustness of our findings, the current study had some limitations. Firstly, although GERD was associated with the risk of insomnia in observational studies [10], the results of MR analysis were not statistically significant, especially the IVW method, which needed further study to explore the causal relationship between GERD and insomnia. Moreover, since the GWAS data used in the study were derived from European populations, it might limit the generalization of the conclusion to non-European populations. It would be helpful to provide more valid conclusions by including more GERD-associated SNPs and a larger sample of more ethnicities. Finally, potential sample overlap might be another source of bias. However, the F-statistics calculated for all SNPs were sufficiently large (ranging from 208 to 669), suggesting that the bias might be minimal [38].

## Conclusions

Our MR study suggests that GERD is an important risk factor for sleep disorders and sleep apnoea, whereas we have found no evidence to support an association between GERD and the risk of insomnia, which may require further study to verify.

## Data Availability

All data produced in the present work are contained in the manuscript/supplemental material.

## Availability of data and materials

The datasets presented in the current study can be found in online repositories.

## Abbreviations

GERD: Gastroesophageal reflux disease
OAS: Obstructive sleep apnoea
RCTs: Randomized controlled trials
MR: Mendelian randomization
SNPs: Single nucleotide polymorphisms
IVs: instrumental variables
GWAS: Genome-wide association study
LD: linkage disequilibrium
IVW: Inverse variance weighted
MR-PRESSO: MR Pleiotropy RESidual Sum and Outlier
CI: Confidence intervals
OR: Odds ratio

## Acknowledgments

The authors acknowledged Jue-Sheng Ong et al. for their contribution, as well as the effort of the FinnGen Consortium in providing high-quality GWAS data for researchers.

## Ethics declarations

### Ethical approval and consent to participate

The GWAS summary data used in this study were obtained from published studies that have been approved by institutional review boards in their respective studies.

### Consent for publication

Not applicable.

### Authors contribution

Study project conception: Guibin Qiao and Zijie Li. Research project organization and execution: Weitao Zhuang and Yong Tang. Data curation: Zijie Li, Junhan Wu, and Haijie Xu. Statistical analysis, design, and execution: Zijie Li and Weitao Zhuang. Statistical analysis review: Guibin Qiao and Yong Tang. Writing of the first draft: Zijie Li, Weitao Zhuang, Junhan Wu, and Haijie Xu. Manuscript review: Guibin Qiao and Yong Tang. Yong Tang and Guibin Qiao take responsibility for the data integrity.

## Supplementary Information

Supplemental Table 1. SNPs used as instrumental variables of GERD on sleep disorders.

Supplemental Table 2. SNPs used as instrumental variables of GERD on sleep apnoea.

Supplemental Table 3. SNPs used as instrumental variables of GERD on insomnia.

Supplemental Figure 1. Leave-one-out analysis of GERD on sleep disorders.

Supplemental Figure 2. Forest plots of each SNPs effect for GERD on sleep disorders and all estimates.

Supplemental Figure 3. Funnel plot of SNPs of GERD on sleep disorders.

