## Supplementary figures and images for "Effect of gastroesophageal reflux disease on sleep disorders: a Mendelian randomization study"

### Supplemental Figure 1

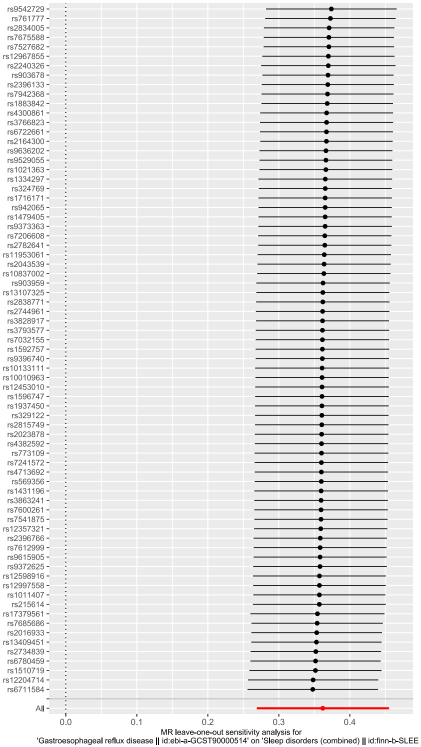


(A) sleep disorders

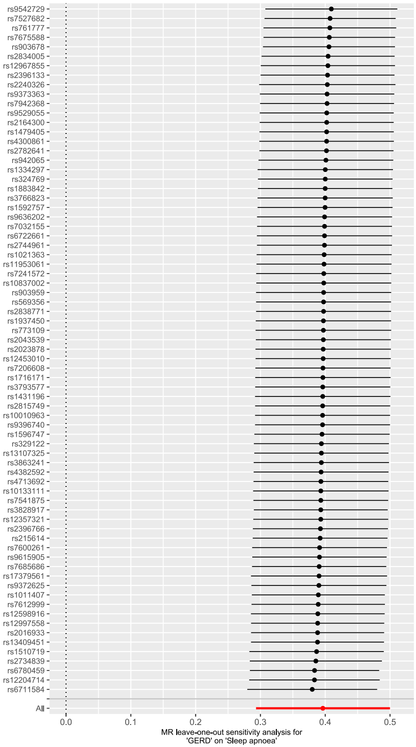


(B) sleep apnoea

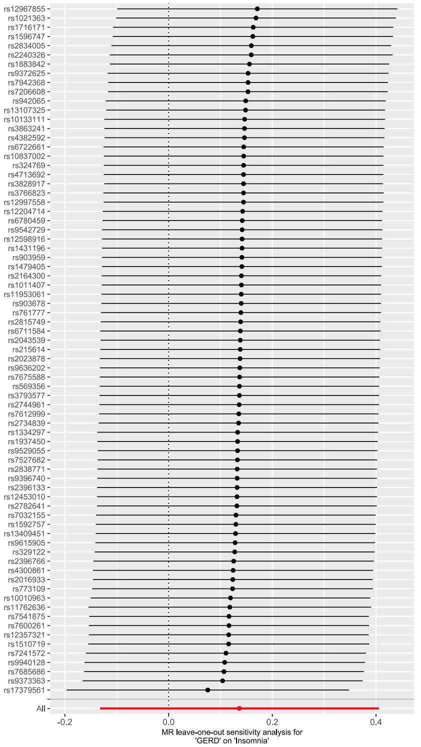


(C) insomnia

### Supplemental Figure 2

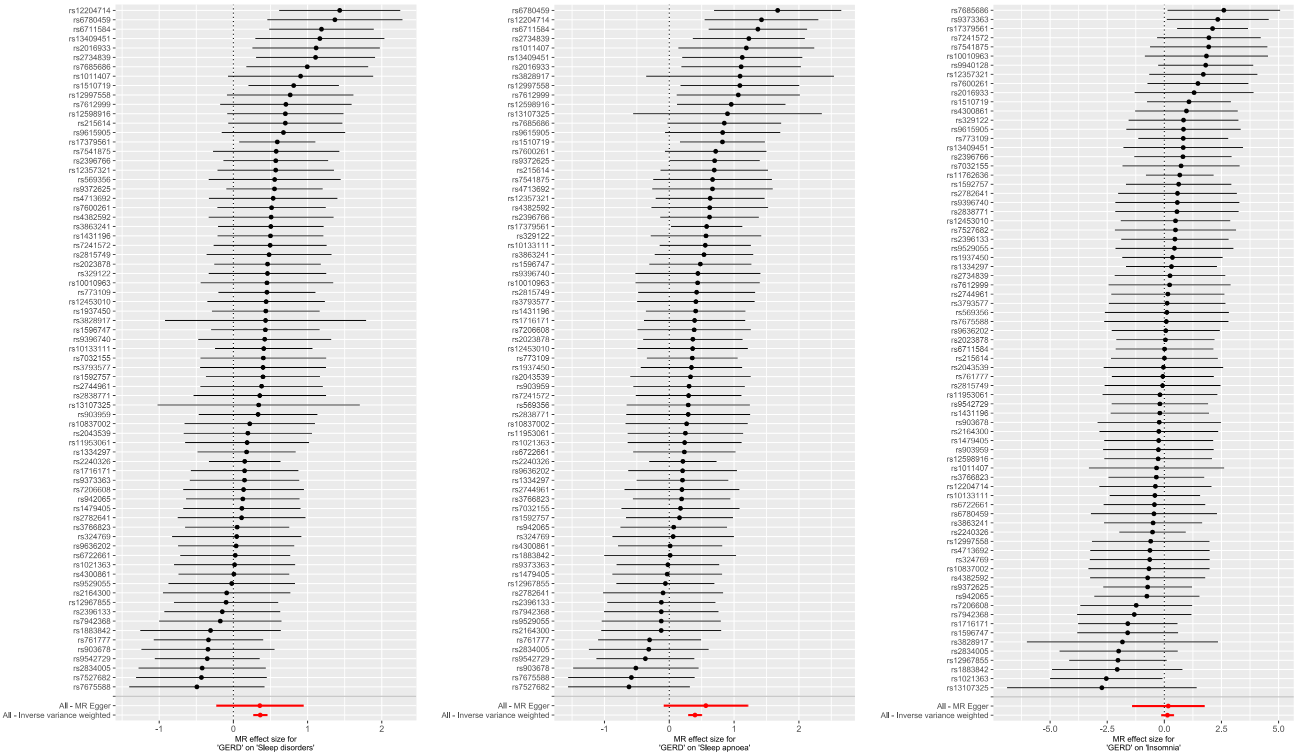


(A) sleep disorders (B) sleep apnoea (C) insomnia

### Supplemental Figure 3

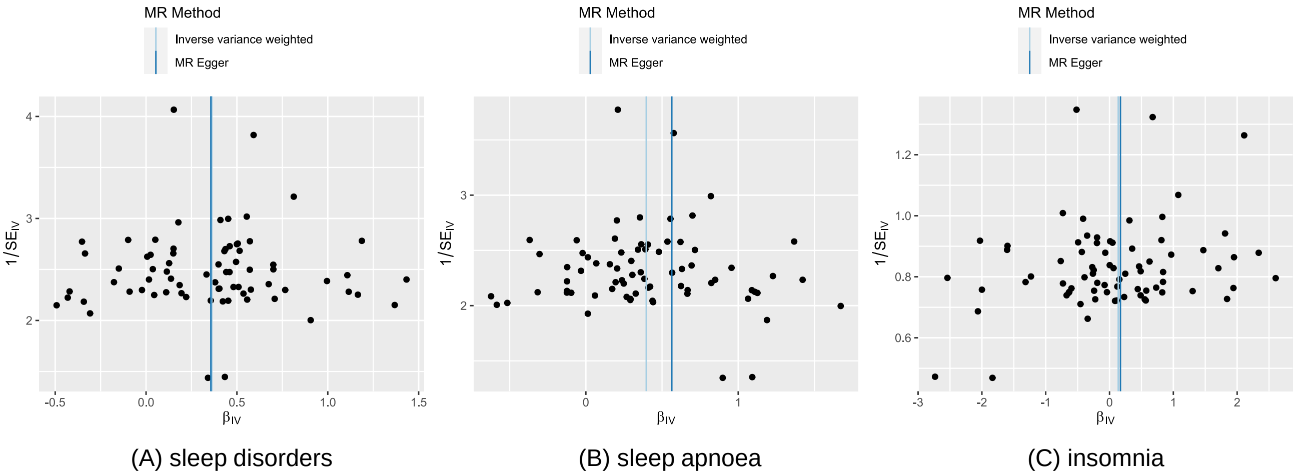
